# Characteristics of COVID-19 fatality cases in East Kalimantan, Indonesia

**DOI:** 10.1101/2020.08.01.20166470

**Authors:** Swandari Paramita, Ronny Isnuwardana, Krispinus Duma, Rahmat Bakhtiar, Muhammad Khairul Nuryanto, Riries Choiru Pramulia Yudia, Evi Fitriany, Meiliati Aminyoto

## Abstract

**Introduction:** Coronavirus Disease (COVID-19) is caused by SARS-CoV-2 infection. On March 2, 2020, Indonesia announced the first confirmed cases of COVID-19 infection. East Kalimantan will play an important role as the new capital of Indonesia. There is attention to the preparedness of East Kalimantan to respond to COVID-19. We report the characteristics of COVID-19 fatality cases in here.

**Methods:** We retrospectively analyzed the fatality cases of COVID-19 patients from the East Kalimantan Health Office information system. All patients were confirmed COVID-19 by RT-PCR examination.

**Results:** By July 31, 2020, 31 fatality cases of patients had been identified as having confirmed COVID-19 in East Kalimantan. The mean age of the patients was 55.1 ± 9.2 years. Most of the patients were men (22 [71.0%]) with age more than 60 years old (14 [45.2%]). Balikpapan has the highest number of COVID-19 fatality cases from all regencies. Hypertension was the most comorbidities in the fatality cases of COVID-19 patients in East Kalimantan.

**Discussion:** Older age and comorbidities still contributed to the fatality cases of COVID-19 patients in East Kalimantan, Indonesia. Hypertension, diabetes, cardiovascular disease, and cerebrovascular disease were underlying conditions for increasing the risk of COVID-19 getting into a serious condition.

**Conclusion:** Active surveillance for people older than 60 years old and having underlying diseases is needed for reducing the case fatality rate of COVID-19 in East Kalimantan.

## Introduction

Coronavirus Disease (COVID-19) is caused by SARS-CoV-2 infection (1). In the past, there have been two epidemics of coronavirus, namely Severe Acute Respiratory Syndrome (SARS) (2), with a mortality rate of 10% (3) and Middle East Respiratory Syndrome (MERS) (4), with a mortality rate of 37% (5).

On March 11, 2020, the World Health Organization (WHO) announced the COVID-19 as a pandemic (6). During March 2020, the new major epidemic center of COVID-19 infection has been identified in Asia, Middle East, Europe, and North America. The number of confirmed cases of COVID-18 outside China had increased rapidly (7). Until July 31, 2020, the USA, Brazil, and India were the countries for the most COVID-19 cases in the world. There are more than 17 million confirmed cases worldwide (8).

On March 2, 2020, President Joko Widodo announced the first confirmed cases of COVID-19 infection in Indonesia (9). On March 18, 2020, the first patient of COVID-19 in East Kalimantan was a patient from Abdul Wahab Sjahranie Hospital Samarinda, the main referral hospital in East Kalimantan for COVID-19 (10). East Kalimantan will play a significant role as the new capital of Indonesia. There is attention to the preparedness of East Kalimantan to respond to COVID-19 (11). As of July 31, 2020, the province has reached 1,426 confirmed cases with 31 deaths (12). Therefore, the study aims to describe the characteristics of fatality cases of COVID-19 patients in East Kalimantan.

## Methods

### Study Design

This study was a descriptive analysis of all fatality cases of COVID-19 patients in East Kalimantan, Indonesia as of the end of July 31, 2020. This study was approved by the Ethical Health Research Commission of Faculty of Medicine Mulawarman University, Samarinda, East Kalimantan, Indonesia. All data were handled to protect patient privacy.

### Data Source

All data collected from COVID-19 fatality case records in the East Kalimantan Health Office information system through July 31, 2020. All cases were included with no sampling was done and no eligibility criteria were used.

### Variables

The database of patients was collected at the time of entry into the East Kalimantan Health Office information system. All confirmed cases were diagnosed based on reverse transcription-polymerase chain reaction (RT-PCR) examination to confirm COVID-19 according to guidelines from the Ministry of Health of the Republic of Indonesia.

### Analysis

For all cases, characteristics of patients were summarized using descriptive statistics. Continuous variables were described as a mean and standard deviation; categorical variables were described as number (%). Statistical analyses were done using Microsoft Excel.

## Results

By July 31, 2020, 31 fatality cases of COVID-19 patients had been identified as having laboratory-confirmed. The highest fatality cases were reaching 24 patients in July 2020. This number has rapidly increased from the last month (Figure 1). Most of the patients were men (22 [71.0%]) with age more than 60 years old (14 [45.2%]). The mean age of the patients was 55.1 ± 9.2 years. Balikpapan has the highest number of COVID-19 fatality cases patients (14 [45.2%]), but Bontang has the highest case fatality rate (4.3%) from all regencies (Table 1). Hypertension was the most comorbidities in the fatality cases of COVID-19 patients in East Kalimantan. Hypertension, diabetes, cardiovascular disease, and cerebrovascular disease were the most common underlying conditions for the fatality cases of COVID-19 patients. There were 9 (29%) patients have more than one comorbidity (Table 2).

**Table 1.** Characteristics of COVID-19 fatality cases.

| Characteristics | Mean ± SD or N (%) |
| --- | --- |
| Age, years | 55.1 ± 9.2 |
| 30-39 years | 2 (6.5%) |
| 40-49 years | 6 (19.4%) |
| 50-59 years | 9 (29.0%) |
| 60-69 years | 13 (41.9%) |
| >69 years | 1 (3.2%) |
| Sex |  |
| Men | 22 (71.0%) |
| Women | 9 (29.0%) |
| Hospital admission of patients |  |
| Province Hospital | 15 (48.4%) |
| Regency Hospital | 9 (29.0%) |
| Private Hospital | 7 (22.6%) |
| Origin of patients |  |
| Balikpapan City | 14 (45.2%) |
| Samarinda City | 11 (35.5%) |
| Paser Regency | 2 (6.5%) |
| Other regencies in East Kalimantan | 4 (12.9%) |
| CFR (Case Fatality Rate) |  |
| East Kalimantan Province | 2.2% |
| Bontang City | 4.4% |
| Samarinda City | 3.6% |
| Balikpapan City | 2.8% |
| Berau Regency | 1.3% |
| East Kutai Regency | 1.0% |
| Kutai Kartanegara Regency | 0.4% |

**Table 2.** Comorbidities of COVID-19 fatality cases.

| Comorbidities |  |
| --- | --- |
| Hypertension | 9 (29.0%) |
| Diabetes | 7 (22.6%) |
| Cardiovascular disease | 6 (19.4%) |
| Cerebrovascular disease | 5 (16.1%) |
| Other comorbidities | 4 (12.9%) |
| Number of comorbidites |  |
| 1 comorbidity | 22 (71.0%) |
| >1 comorbidities | 9 (29.0%) |

**Figure 1.**
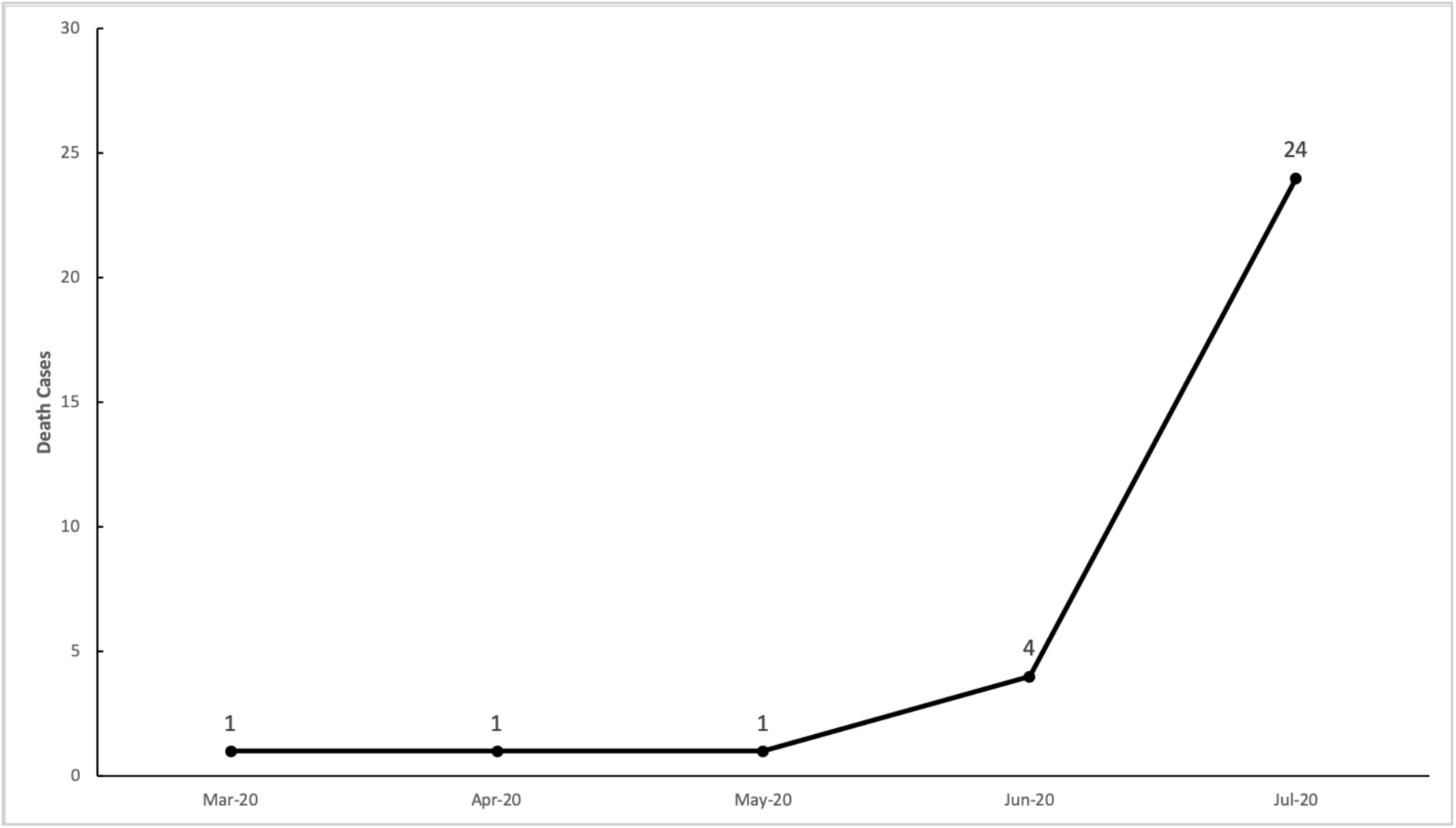
The number of COVID-19 fatality cases in East Kalimantan (March to July 2020)

## Discussion

This study reports the characteristics of 31 fatality cases of COVID-19 patients in East Kalimantan, Indonesia. As of July 31, 2020, there have been 1,426 COVID-19 positive cases in East Kalimantan (12). On that date, there are 108,376 positive confirmed cases of COVID-19, with 65,907 people recovering and 5,131 patients have died in Indonesia (13).

The progress of COVID-19 cases in March 2020 in East Kalimantan showed a total case fatality rate (CFR) of 1.85% (11). There was only one fatality case of COVID-19 patients in Samarinda in June 2020 (14). But there were already 10 fatality cases in Samarinda in July 2020. The WHO estimates that 14% of infected cases are severe and require hospitalization. Five percent of infected cases are very severe and require intensive care admission, mostly for ventilation with 4% of the infected die. COVID-19 pandemic may cause the death of 6% of the global population (15).

The mean age of the dying patients in this study was 55.1 ± 9.2 years and 14 (45.2%) fatality case patients with age more than 60 years old. These results are similar to the previous retrospective cohort study in Indonesia. Data from the majority of the fatality cases, older people were associated with an increased risk of death (16). This is consistent with a study about the mortality rate of COVID-19 with a median age of 56.0 years. In-hospital death was associated with older age on admission (17).

The majority of fatality cases of COVID-19 patients in East Kalimantan are male (22 [71.0%]). This is consistent with a meta-analysis study which showed that men took the largest percentage in the distribution of COVID-19 according to gender (18). These results are similar to the previous study at Abdul Wahab Sjahranie Hospital Samarinda which showed that most patients were also male (10).

Hospital admission of 15 (48.4%) fatality case patients were in province hospital. There are 16 main referral hospitals for COVID-19 in East Kalimantan. Abdul Wahab Sjahranie Hospital in Samarinda and Kanujoso Djatiwibowo Hospital in Balikpapan are two of province hospitals in East Kalimantan. Province hospitals have more medical equipment needed for COVID-19 patients in critical condition, compared with regency or private hospitals (12).

The highest number of COVID-19 fatality case patients (14 [45.2%]) were in Balikpapan, but Bontang has the highest case fatality rate (4.3%) from all regencies. East Kalimantan has been assigned as the new capital of the Republic of Indonesia. The flow of people who came in and out of this province has increased recently. Balikpapan, Samarinda, and Bontang are three regencies with the highest population density in East Kalimantan (11). The spread of COVID-19 depends on high population densities. Controlling infection rates is key to COVID-19 control and it depends on population densities (19).

Hypertension, diabetes, cardiovascular disease, and cerebrovascular disease were the most common underlying conditions for the fatality case of COVID-19 patients. There were 9 (29%) patients have more than one comorbidity. These results are similar to systematic review and meta-analysis study which showed that comorbidities are risk factors for severe COVID-19 patients. The most prevalent comorbidities were hypertension and diabetes, followed by cardiovascular disease (20). This is also consistent with a meta-analysis study which showed that hypertension, diabetes, cardiovascular disease, and cerebrovascular disease were risk factors associated with COVID-19 patients (21). These results are similar to the previous study at Wuhan, China which showed that underlying hypertension was significantly associated with severe COVID-19 on hospital admission. Hyperglycemia in diabetes was associated with death in patients with severe COVID-19 in Wuhan, China (22).

## Conclusion

Patients with older age and comorbidities need observation and early intervention to prevent the development of serious COVID-19 conditions. Active surveillance for people older than 60 years old and having underlying diseases is needed for reducing the case fatality rate of COVID-19 in East Kalimantan.

## Data Availability

The datasets generated during and/or analyzed during the current study are available from the corresponding author on reasonable request.

